# Protocol for semantic segmentation of spinal endoscopic instruments and anatomic structures : how far is robotic endoscopy surgery?

**DOI:** 10.1101/2024.04.14.24305785

**Authors:** Guoxin Fan, Guanghui Yue, Zhouyang Hu, Zhipeng Xu, Jianjin Zhang, Hong Wang, Xiang Liao

## Abstract

**Background:** Automatic analysis of endoscopic images will played an important role in the future spine robotic surgery. The study is designed as a translational study to develop AI models of semantic segmentation for spinal endoscopic instruments and anatomic structures. The aim is to provide the visual understanding basis of endoscopic images for future intelligent robotic surgery.

**Methods:** An estimate of 500 cases of endoscopic video will be included in the study. More data may also be included from the internet for external validation. Video clip containing typical spinal endoscopic instruments and distinct anatomic structures will be extracted. Typical spinal endoscopic instruments will include forceps, bipolar electrocoagulation, drill and so on. Endoscopic anatomic structures will include ligament, upper lamina, lower lamina, nerve root, disc, adipofascia, etc. The ratio of training, validation and testing set of included samples is initially set as 8: 1: 1. State-of-art algorithm (namely UNet, Swin-UNet, DeepLab-V3, etc) and self-developed deep learning algorithm will be used to develop the semantic segmentation models. Dice coefficient (DC), Hausdorff distance (HD), and mean surface distance (MSD) will be used to assess the segmentation performance.

**Discussions:** This protocol firstly proposed the research plans to develop deep learning models to achieve multi-task semantic segmentation of spinal endoscopy images. Automatically recognizing and simultaneously contouring the surgical instruments and anatomic structures will teach the robot understand the surgical procedures of human surgeons. The research results and the annotated data will be disclosed and published in the near future.

**Metadata:** The authors did not receive any funding for this work yet.

The authors have declared no competing interests.

No data analyzed during the current study. All pertinent data from this study will be disclosed upon study completion.

## Introduction and rationale

Spinal endoscopic surgery (SES) is one of the most popular minimally invasive technique to treat degenerative spinal disease(1). SES is deemed as a prior surgical option over open spine surgery for single-level disc herniation, spinal stenosis and even interbody fusion, as SES has merits of less trauma, minimal blood loss and fast recovery(2). However, SES is associated with longer operation time and more radiation exposure, which can be minimized by detailed preoperative planning and intelligent intraoperative assistance(3). Thus, there is a great needed to introduce intelligent robotic technique to optimize SES(4).

The current robotic surgery has already played an important role in preoperative planning, intraoperative calibration and navigation (5). Spine-related robotic surgery mainly focused on spinal fusion and instrumentation procedures, and others have tried some complicate tasks such as vertebroplasties, tumor resections and ablations, deformity correction(6), and stabilizing tubular retractors(7). The future direction of spine robotic surgery may include decompression, discectomy or mobilization of neural elements (8, 9). However, it should also be noted that automated analysis of endoscopic images will played an important role in the future spine robotic surgery. Thus, this protocol aimed to deploy automated image analysis of SES with artificial intelligent (AI) technique.

## Methods

The study is designed as a translational study to develop AI models of semantic segmentation for spinal endoscopic instruments and anatomic structures. The aim is to provide the visual understanding basis of endoscopic images for future intelligent robotic surgery.

### Data collection

An approximately 3-year case-series data will be retrospectively collected from Huazhong University of Science and Technology Union Shenzhen Hospital, Shenzhen, China. This study will adhere to the Declaration of Helsinki, and ethics have been approved by the ethics committee. Ethics committee of Huazhong University of Science and Technology Union Shenzhen Hospital has waived the ethical approval for this work, as the study did not include any identifiable personal data.

### Inclusion criteria

Spinal endoscopic video of lumbar decompression, cervical decompression, lumbar discectomy, and cervical discectomy.

### Exclusion criteria

Patients receive prior lumbar or cervical surgery of decompression or discectomy at the same level.

### Sample size

An estimate of 500 cases of endoscopic video will be included in the study. More data may also be included from the internet for external validation.

### Data annotation

Video clip containing typical spinal endoscopic instruments and distinct anatomic structures will be extracted. Typical spinal endoscopic instruments will include forceps, bipolar electrocoagulation, drill and so on (**Figure 1**). Endoscopic anatomic structures will include ligament, upper lamina, lower lamina, nerve root, disc, adipofascia, etc (**Figure 2**). All annotations were labeled by one independent expert using Pair V 2.9 and then revised by another two experts. Any disagreement would be voted by the three experts and the final segmented masks were regarded as ground truth. In addition to record the information of different surgery for each sample, we will also mark the samples whether they come from the large channel (a diameter of 11.0mm) or the small channel (a diameter of 7.5mm), and different brand for surgical instruments.

**Figure 1.**
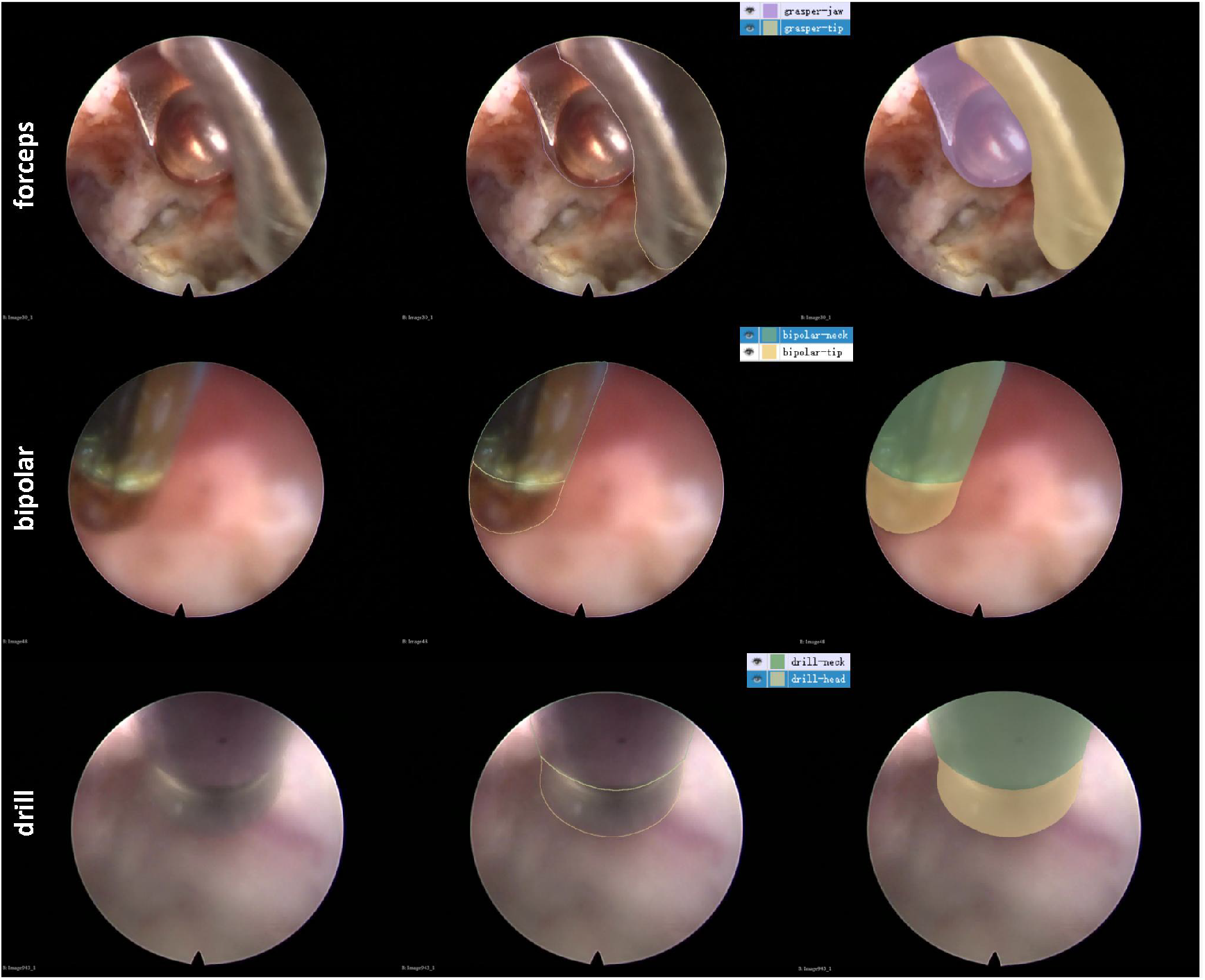
Annotations of typical surgical instruments under endoscopy.

**Figure 2.**
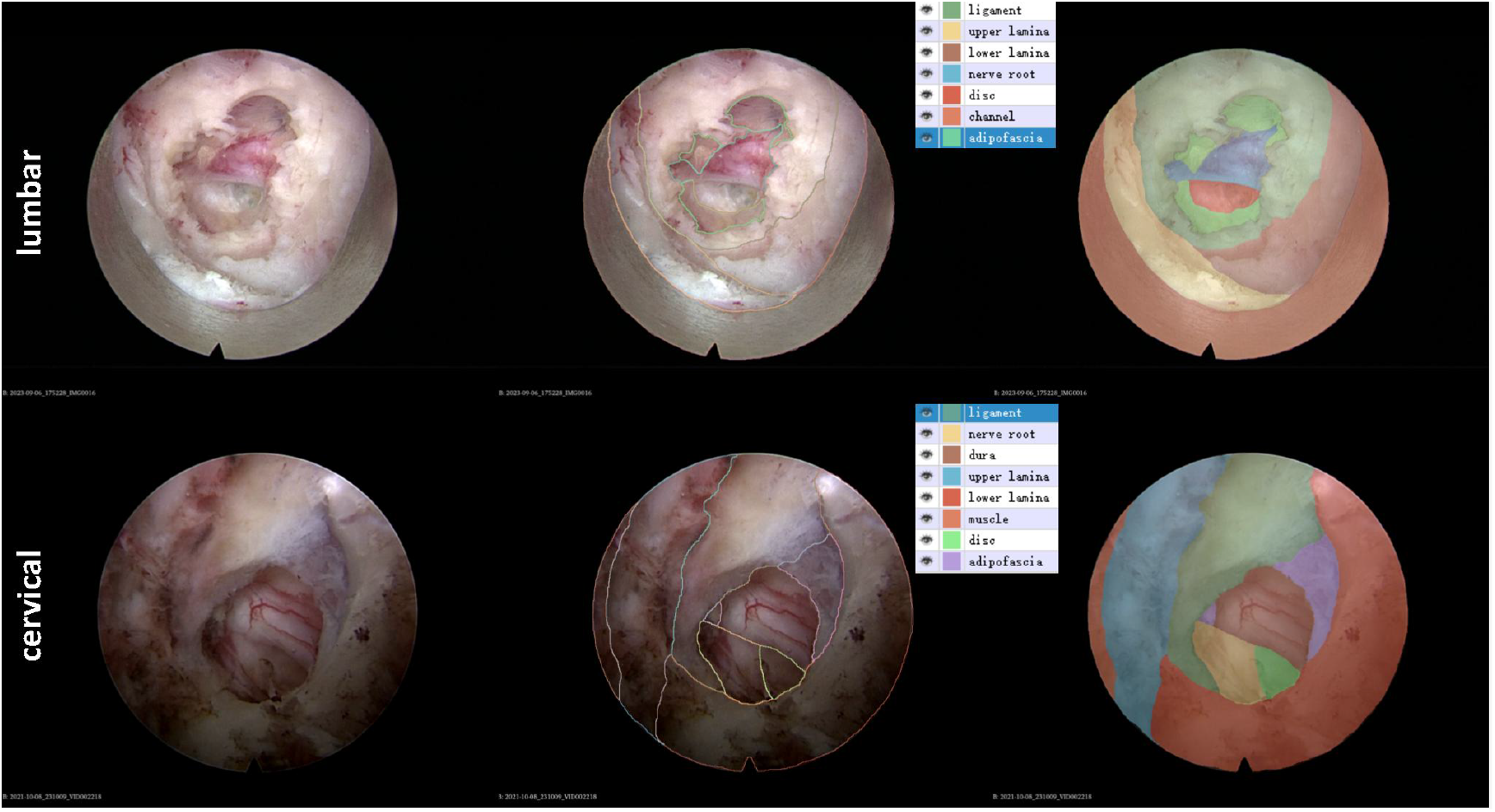
Annotations of anatomic structures under endoscopy.

### Development of AI models

The ratio of training, validation and testing set of included samples is initially set as 8: 1: 1. State-of-art algorithm (namely UNet, Swin-UNet, DeepLab-V3, etc) and self-developed deep learning algorithm will be used to develop the sementic segmentation models with Python 3.9. Five-fold cross-validation and external validation will be used to validate the deep-learning models.

### Outcome measures

Dice coefficient (DC), Hausdorff distance (HD), and mean surface distance (MSD) will be used to assess the segmentation performance.

## Discussion

Robotic surgery should raise interest in automated surgical image analysis(10), but the automated image analysis of intraoperative SES is scarce. This protocol takes the lead in envisioning the directions of robotic SES and proposed the specific research plans of semantic segmentation of spinal endoscopic images. The research results and the annotated data will be disclosed and published in the near future.

Robotic SES may include three stages or directions, namely assistant-robot phase, telerobotics phase, and automated-robot phase (**Figure 3**). During assistant-robot phase, the surgical robot may play an assistant role in holding the endoscopic camera, which will automatically adjust the depth and the angle of the endoscopic camera. Additionally, the assistant robot is expected to facilitate the fluoroscopy, localize the bleeding spot, quantify the decompression area, etc. During telerobotics phase, surgeons may not need to scrub in the surgery, but focus on manipulate the surgical robot. In addition to the above mentioned assistance, the surgical telerobot is also expected to complete the operation while the manipulated surgeons are thousands of miles away. During automated-robot phase, the surgical robot is expected to make preoperative surgical plans and conduct the operation on its own, while the surgeons are far more like the supervisors. These scenes are not far away, while humanoid robots have already come into our life and doing housework on their own(11).

**Figure 3.**
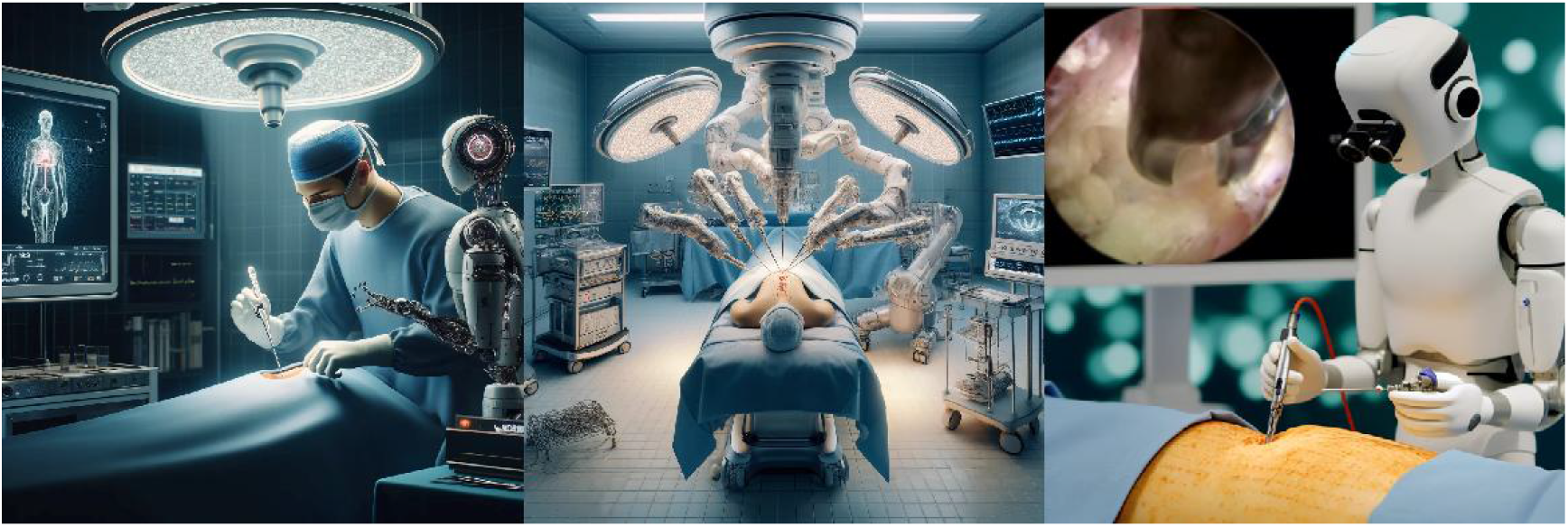
Three kinds of robotic SES generated by DALL·E 3 model.

The recent development of deep learning algorithms has enable and accelerated the advent of robot time. The main reason why the development of surgical robot falls behind the housework robot is the lack of annotated data. For spine surgical robots, the endoscopy images provide the opportunity to train the robots with accessible, standardized and massive data. However, very few studies have ever noticed the importance of automated image analysis of intraoperative SES. One study endeavored to developed computer-assisted system with deep learning and spinal endoscopy images in order to assist the endoscopists in recognizing the neural element(12). Other study focused on automatically localizing the tip of the surgical instruments for biportal endoscopic spine surgery(13). To the best of our acknowledgement, these were the only two studies tried to develop automatic system with spinal endoscopy images. No previous studies have ever noted the importance of semantic segmentation of spinal endoscopy images for robotic SES.

This protocol firstly proposed the research plans to develop deep learning models to achieve multi-task semantic segmentation of spinal endoscopy images. The segmentation targets include various typical surgical instruments and all kinds of anatomic structures during SES. Automatically recognizing and simultaneously contouring the surgical instruments and anatomic structures will teach the robot understand the surgical procedures of human surgeons. For instance, the drill is usually used to remove the bones or grind the calcified disc. The surgical robot not only need to know when and how to use the drill, but also need to localize the drill and quantify the decompression area. Therefore, all this kinds of complex tasks rely on the semantic segmentation of various typical surgical instruments and all kinds of anatomic structures. Yet, it should be also noted that this protocol did not include the video data of endoscopic fusion procedure, which will be explored in the future.

## Authors’ Contributions

Guoxin Fan, MD: Conceptualization, data curation, draft writing.

Guanghui Yue, PhD: methodology, validation, formal analysis

Zhouyang Hu, PhD: manuscript preparation

Zhipeng Xu, MBBS: figure preparation.

Jianjin Zhang, MBBS: data annotations.

Hong Wang, MBBS: data annotations.

Xiang Liao, MD; Conceptualization, project administration

